# Brain morphometry and psychomotor development in children with PCH2A

**DOI:** 10.1101/2024.12.09.24318534

**Authors:** Pablo Pretzel, Antonia Herrmann, Alice Kuhn, Anna-Lena Klauser, Julia Matilainen, Elias Kellner, Maren Hackenberg, Simone Mayer, Lucia Laugwitz, Markus Uhl, Samuel Groeschel, Wibke Janzarik

## Abstract

**Introduction:** Pontocerebellar hypoplasia type 2A (PCH2A) is a rare neurogenetic disease causing severe cognitive and motor impairment. We report on brain morphometry and psychomotor development of affected children.

**Materials and Methods:** We analyzed 78 cerebral MRI datasets of 57 patients with genetically proven PCH2A. Volumetry and in-plane measurements were performed in cerebellum, neocortex and pons. Supratentorial width and width of the anterior horns of the lateral ventricles was used to calculate the Evans index.

Caregivers of 65 patients with PCH2A (7 months to 33 years) filled in a survey assessing motor and cognitive development. Developmental status was compared to MRI measurements.

**Results:** In children with PCH2A, cerebellar volume was markedly smaller than in healthy children at birth, with slower increase and stagnation at around 12 months. No cerebellar growth was observed in the cranio-caudal axis. Long-term data did not show a decrease in cerebellar volume or in-plane measurements. Supratentorial measurements showed progressive microcephaly and a continuous increase of the Evans index, indicating progressive cerebral atrophy. Patients showed severely impaired cognitive and motor development. Developmental regression was reported only for a minority. No statistical relationship between brain measurements and cognitive or motor development was observed.

**Conclusion:** MRI of patients with PCH2A shows limited cerebellar growth during infancy, especially restricted along the cranio-caudal axis. After infancy, cerebellar volume remains relatively stable. Supratentorial measurements indicate slowly progressive atrophy. Psychomotor development is severely impaired, but regression is rare.

**Highlights:**

- MRI (57 patients) and developmental data (65 patients) of children with PCH2A
- Cerebellar growth is affected in a spatial manner, resulting in the dragonfly sign
- Ongoing supratentorial atrophy, potentially secondary to cerebellar pathology
- Psychomotor development is delayed, but takes place and regression is rare
- No relation between brain measurements and developmental trajectories

## Introduction

Pontocerebellar hypoplasia (PCH) is a group of very rare, autosomal recessive genetic diseases. In the present study, we focus on PCH type 2A (PCH2A), the most common subtype (Rudnik-Schöneborn et al, 2014). This subtype is defined by a specific homozygous pathogenic variant p.A307S (c.919G>T) in the TSEN54 gene, which is suggested to result in impaired tRNA processing (Kasher et al, 2011; Namavar et al, 2011a), and potentially, mRNA decay (Paushkin et al, 2004; Hurtig et al, 2021).

Children with PCH2A develop symptoms already in early infancy, such as impaired swallowing, restlessness, and a choreatic movement disorder. Additional neurological symptoms, such as long-lasting dystonic attacks, spasticity, and epileptic seizures, often develop within the first years of life (Sánchez-Albisua et al, 2014). Both motor and cognitive development are severely impaired. All children with PCH2A show severe progressive microcephaly (Kuhn et al, 2024). Life expectancy is limited, and many affected patients die during childhood and adolescence (Rudnik-Schöneborn et al, 2014, Sánchez-Albisua et al, 2014; Kuhn et al, 2024).

Neuroimaging of children with PCH2A reveals eponymous, severe hypoplasia of pons and cerebellum, with a relative sparing of the cerebellar vermis, resulting in the characteristic “dragonfly pattern” (Namavar et al, 2011b). Histological findings indicate loss of cerebellar cortex, fragmentation of dentate nuclei, and near absence of transverse pontine fibers (Barth et al, 2007; Rudnik-Schöneborn et al, 2014). Supratentorial structures are also affected, with reduced cerebral volume and enlarged ventricles (Ekert et al, 2016).

However, due to the rarity of PCH2A, little data is available on the detailed longitudinal trajectory of brain measurements of affected children before and beyond the first year of life. Only a small number of prior studies provided cross-sectional data, mainly from the first months of life: van Dijk et al (2021) reported on 18 patients aged 14.8 months or less, Ekert et al (2016) reported on 24 children aged 17 years or less (median 10 months).

In the present study, we aimed to broaden the understanding of the brain structural phenotype and developmental phenotype of PCH2A by investigating a larger cohort, with data including repeated MRI and MRI of older patients. Specifically, this approach would allow to characterize the long-term trajectory of the disease and assess whether progressive developmental regression or cerebral atrophy occur with increasing age. A comprehensive characterization of the course of PCH2A will benefit clinicians, researchers, and affected patients.

## Materials & Methods

Patients with PCH2A were recruited via the German patients` organization PCH-Familie e.V. and included data from the first natural history study (Sánchez-Albisua et al, 2014). Inclusion criteria were genetic confirmation of the homozygous pathogenic variant p.A307S of the *TSEN54* gene and written informed consent of the legal guardian. The study was approved by the ethics committee of the University of Freiburg, Germany (decision no. 20-1040) and by the ethics committee of the University of Tübingen (decision no. 105/2012BO2), as part of a natural history study on patients with PCH2A.

Parents completed a disease-specific survey with data on motor and cognitive development. Additionally, available imaging data and medical records were collected for a retrospective analysis, if available. In a few cases, parents did not complete the parent survey, but gave consent to use MRI data for analysis.

### Brain morphometry

In MRI datasets of sufficient quality (effective slice thickness ≤ 3mm, field of view spanning the entire brain), volumes of cerebellum, pons, supratentorial structures, and of the ventricles were measured by manually masking the respective structures and calculating the volume of the resulting masks (see Supplementary Figure 1). Additionally, in-plane size measurements of specific brain structures were taken in all MRI datasets with a suitable field of view (see Supplementary Figure 2): Infratentorial measurements included cerebellar transverse diameter, anterior-posterior length, superior-inferior height (averaged between left and right lobe), and midline anterior-posterior diameter of the pons. Supratentorial measurements comprised the largest transverse diameter of the anterior horns of the lateral ventricles, as well as the transverse diameter of the cerebrum in the same axial plane. From these, the Evans index was calculated to assess supratentorial brain atrophy (Sari et al, 2015). All in-plane measurements of size dimensions were performed using DeepUnity (Dedalus Healthcare Group, Bonn, Germany). Masks for volumetry were created using Nora (nora-imaging.com).

Reference data of cerebellar and cerebral volume were applied from a previous study with similar methodological approach (Ekert et al, 2016). Reference data of pontine diameter were taken from the literature (Promnitz et al, 2022). To assess the reliability of the measurements conducted by the first rater (AH), both, in-plane as well as volumetric measurements were independently repeated in 34 and 10 randomly selected datasets, respectively, by a second researcher (PP), blinded to the original results. Inter-rater variability was assessed using the Intraclass Correlation Coefficient (Koo and Li, 2016). Statistical analyses were performed using SPSS (IBM, Armonk, NY) and R (R core team, 2017), and visualized using SPSS and ggplot (Wickham, 2009).

### Assessment and classification of psychomotor development

As part of the parent survey, the developmental history of the participating patients with PHC2A was assessed over a range of skills across both the motor and cognitive domain, as described in a previous publication (Sánchez-Albisua et al, 2014). For each skill, parents were asked whether and at what age their child had reached the respective skill, and whether development of this skill was still in progress, or at what age it had stagnated. Additionally, parents were asked to assess, whether the child showed the respective skill regularly or only intermittently, or whether it had unlearned the skill at some point. Parental answers from the survey were cross-validated with the medical records, and inconsistencies were discussed in an additional telephone interview with the parent. In the rare instances of persistent discrepancies between parental assessment and documented clinical findings, we accepted the parental assessment, considering the more continuous observation of the affected child by the parents.

To categorize the variability of psychomotor development in children with PCH2, for each skill, the median age at achievement was calculated among all children who had ever demonstrated the skill.

Based on these cohort-specific development timelines, we calculated for each child the ratio of skills it had achieved to those typically achieved by children with PCH at that age. The 15 % of children with the highest ratio were classified as above-average development, while the 15 % with the lowest ratio were classified as below-average development. The remaining children were classified as average development. We then compared cerebellar and supratentorial measurements against these developmental classifications.

## Results

72 patients with genetically proven PCH2A were enrolled in the study. For 57 of these, a total of 78 MRI datasets of sufficient quality for in-plane measurements were available, three of which were obtained in-utero. Of these datasets, 48 were of suitable quality for volumetry (effective slice thickness ≤ 3mm). From 17 patients, two imaging datasets were available, and from 2 patients, three longitudinal data points were available. No imaging data was available for 15 patients. Due to insufficient quality, 3 MRI datasets were excluded from the analyses. Developmental data was available in 65 patients with PCH2A, while for 7 children only MRI was provided. Cohort details are listed in Table 1.

**Table 1:** Cohort details.

|  |  |
| --- | --- |
| Total enrolled patients | 72 |
| Patients with available MRI | 57 |
| Total available MRI datasets | 78 |
| MRI datasets suitable for volumetry | 48 |
| Gender of patients with available MRI | 33 female (58 %), 24 male (42 %) |
| Median age at MRI | 4,5 months (31 weeks of gestation – 18.7 years) |
| Patients with developmental data | 65 |
| Gender of patients with developmental data | 33 female (51 %), 32 male (49 %) |
| Median age at time of the survey | 6,3 years (7 months – 33 years) |

### Volumetric measurements

Results of the volumetric measurements are illustrated in Figure 1. Already at birth, a stark difference in cerebellar volume was observed between children with PCH2A and healthy controls (Figure 1, upper left). Patients then showed limited cerebellar growth during the first months of life, after which the interpolated logarithmic growth curve flattened progressively and plateaued at a volume of approximately 20 ml (upper right). Healthy controls showed a markedly faster and ongoing cerebellar growth. There was no difference between male and female patients.

**Figure 1:**
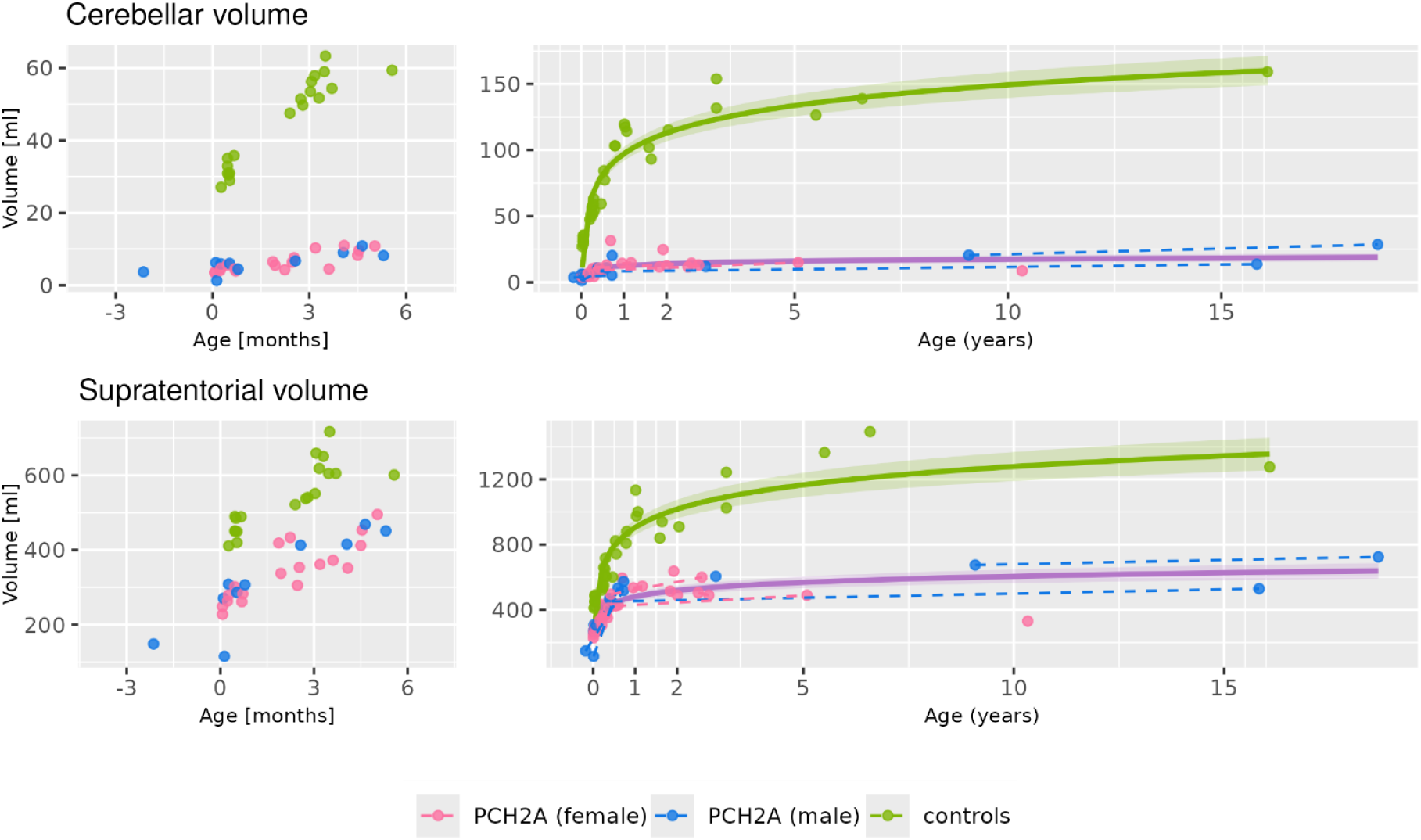
Volumetry of the cerebellum (top) and supratentorial structures (bottom). Volumes of both structures show markedly decreased brain growth in children with PCH2A as compared to healthy controls. Growth restriction is more pronounced in the cerebellum than in supratentorial brain structures, especially during the first months of life (left). After the first year, brain volumes remain stable in patients with PCH2A, whereas healthy controls show continuous growth of brain structures during childhood (right). Measurements from longitudinal data of the same patients (dashed lines) fit to the interpolated logarithmic curve (solid line, lighter shade: 95% confidence interval). Values on the left side of time point zero reflect prenatal MRI. No differences are observed between male and female patients.

Differences in supratentorial brain volume were less pronounced at birth, and supratentorial volume in patients appeared to lag behind healthy controls only moderately during the first months of life (Figure 1, bottom left). After the first year of life, however, supratentorial cerebral growth also stagnates in patients with PCH2A, and remains at markedly lower volumes than in healthy controls (bottom right). Again, no differences were observed between male and female patients.

Both for cerebellar and supratentorial volume, individual data points and longitudinal follow-up data do not show a progressive loss of volume. Longitudinal follow-up data show a good fit to the interpolated logarithmic growth curve.

### Measurements of cerebellar and pontine diameters over time

Measurements of cerebellar and pontine diameters complement the volumetric approach (Figure 2).

**Figure 2:**
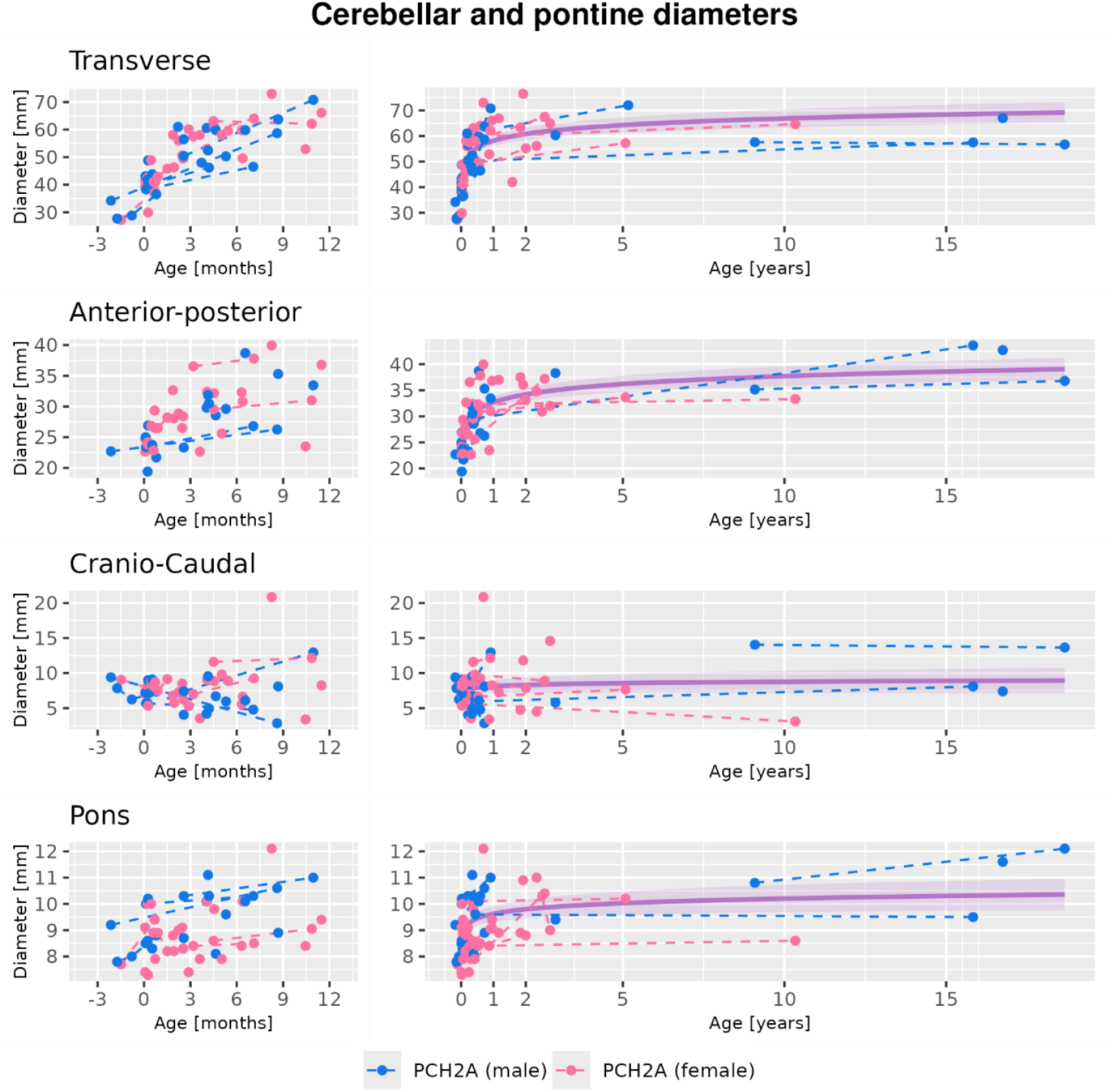
Cerebellar measurements in transversal (top), anterior-posterior (second row) and cranio-caudal (third row) direction, as well as anterior-posterior pontine diameter (bottom row). Over the first year of life, cerebellar size increases in transverse diameter and anterior-posterior length, but not in cranio-caudal height (left). Together, this results in the characteristic dragonfly pattern commonly observed in PCH2A. After the first years of life, cerebellar growth ceases in all dimensions (right). Pontine diameter increases slightly in the first year of life, then also stagnates. Dashed lines indicate longitudinal data of the same patient and provide a good fit to the interpolated logarithmic curves (solid lines). No clear growth difference is observed between male and female patients. Values in the negative range reflect prenatal MRI (3 cases) or MRI of a premature child (1 case). See Supplementary Figure 2 for an illustration of measurements.

The growth curves for transverse cerebellar diameter (top row) and anterior-posterior diameter of the cerebellum (second row) follow a similar pattern to cerebellar volumetry, with clear growth in children with PCH2A in the first months, but at a slower pace and earlier stagnation of growth than in healthy controls. A similar pattern is also observed in pontine anterior-posterior diameter (bottom row). Cranio-caudal diameter of the cerebellar hemispheres, however, remains relatively stable after birth and does not show any growth, neither during infancy nor at a later developmental stage (Figure 2, third row). In some children with follow-up MRI, we observed a slight decrease of cranio-caudal cerebellar diameter, specifically noticeable in one child with 9,5mm diameter immediately after birth to 3mm only a few months later (see Supplementary Figure 3). Overall, longitudinal MRI data from individual patients showed a good fit to the interpolated growth curves. No significant difference in diameters was observed between male and female subjects.

### Measurements of supratentorial diameter over time

Supratentorial in-plane measurements are illustrated in Figure 3. Transverse cerebral width (top row) essentially shows a pattern of growth within the first few years and subsequent stagnation similar to cerebellar structures, but at a later age. At the same time, however, lateral ventricle width shows a sustained increase (middle). Together, these two growth tendencies lead to a continuous increase of the Evans index (bottom), which, shortly after birth, surpasses the value of 0.3 defined as the upper threshold for healthy controls independent of age (Sari et al, 2015). No MRI showed signs of elevated intracranial pressure or obstructed circulation of cerebrospinal fluid.

**Figure 3:**
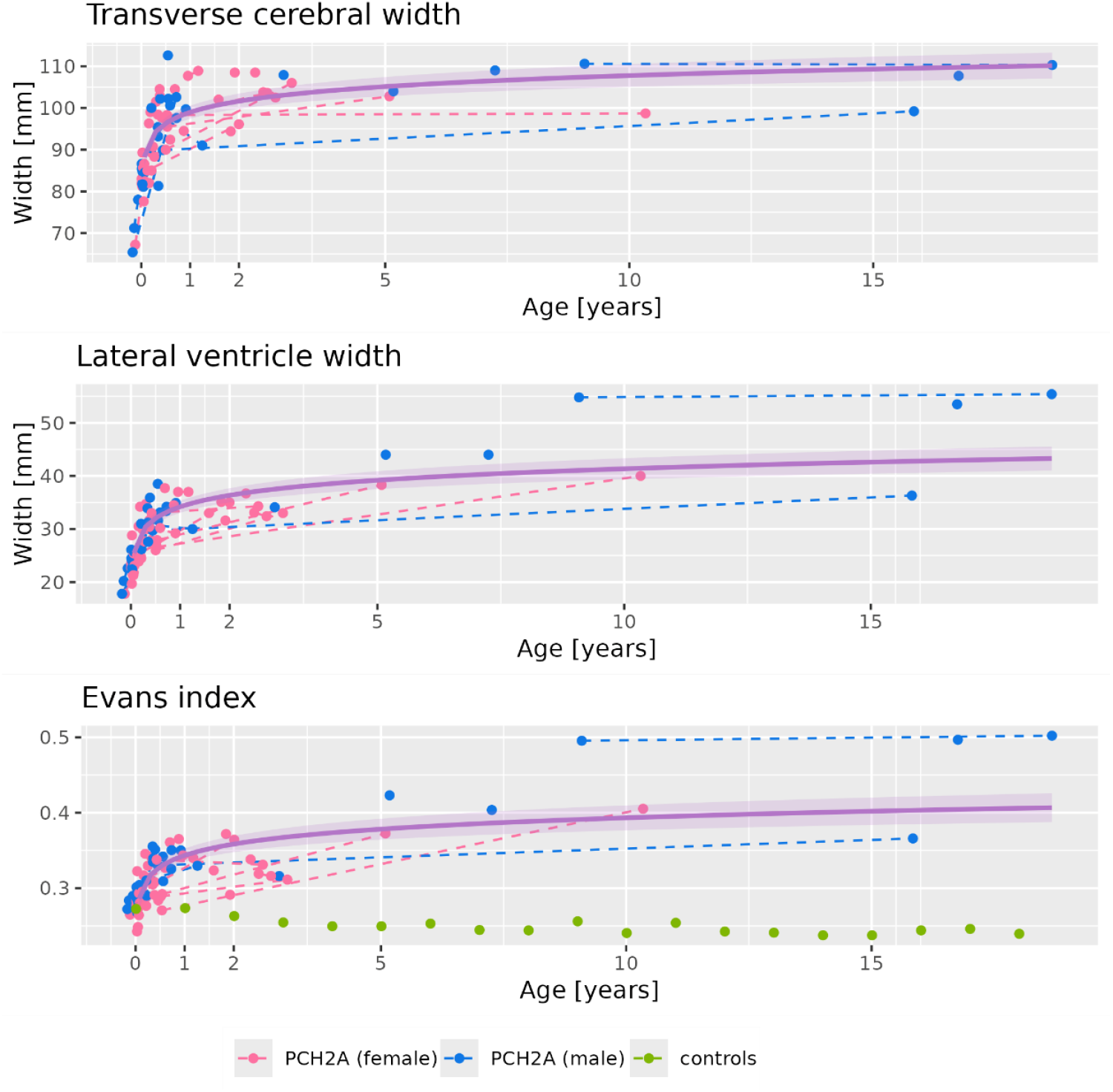
Supratentorial measurements: Transverse cerebral width (top), width of the anterior horns of the lateral ventricles (middle), Evans index (bottom). While transverse cerebral width stagnates around the age of 12 months, similar to infratentorial measures, the width of the anterior horns of the lateral ventricles continues to increase, causing the Evan’s index to rise above the threshold of 0.3 established for healthy children.

### Inter-rater correlation

The intraclass correlation coefficient (ICC) for the volumetric measurements was 0.97 for cerebellar measurements (95 confidence-interval [CI] 0.88-0.99) and 0.991 for supratentorial volumes (95 % CI 0.67-1.0). For the two-dimensional diameters the ICC was between 0.92 and 0.97 for the three cerebellar diameters (CI 0.82-0.96 and 0.93-0.98) and 0.86 for Pons (CI 0.71-0.94). The ICC for supratentorial measurements was 0.94 (CI 0.88-0.97) for the cerebral diameter and 0.99 for the diameter of the lateral ventricles (CI 0.97-0.99).

### Psychomotor development

Development of motor skills is illustrated in Figure 4 (top). Basal motor skills were achieved by a considerable number of children, such as head control (68 % of all children older than the median age at which the skill was achieved) or grasping attempts (74 %). Rolling over and targeted grasping was learned by 52 % and 40 %, respectively. More advanced motor skills, however, were rarely achieved (crawling: 16 %, on all fours: 12 %, unsupported sitting: 9 %). Overall, all skills were acquired with a significant delay in comparison to healthy children. 10 children were reported to never have learned any of the queried motor skills. For all motor skills, it was indicated for at least one child that the skill was later unlearned (grasping attempts and rolling over: 5 children each, all other skills: 1-2 children each).

**Figure 4:**
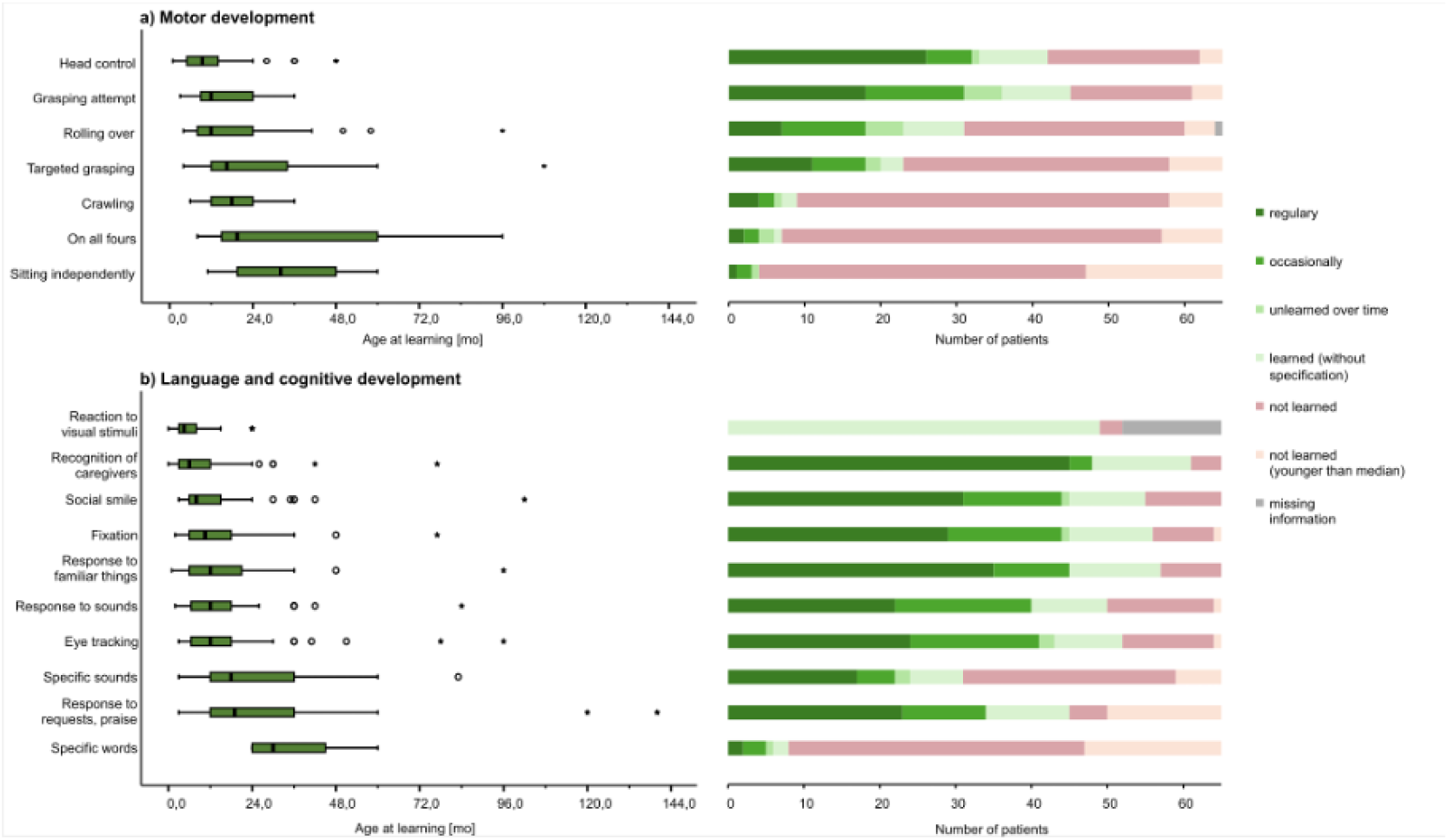
Motor development (top) and language and cognitive development (bottom) of children with PCH2A, as reported by caregivers. Left side: Age at which the skill was first demonstrated. Right side: Absolute number of children that achieved, unlearned or never achieved the respective skill. Total green bar: Number of children that ever achieved the skill. Individual shades of green differentiate the frequency with which the skill was demonstrated. Red bar: Number of children that never achieved the skill. Light orange: Children who were younger at the time of assessment than the median age at which the skill was achieved by children with PCH2A. Motor development (top): Simpler skills like head control, turning and grasping attempts were demonstrated by about half the children, mostly within the first few years of life, but considerably later than in healthy children. More complex abilities were only demonstrated by a small number of children, and at a higher and more widely distributed age. For each skill, a number of parents reported that their child had achieved the skill at some point but did not exhibit it anymore. Cognitive development (bottom): A considerable number of children started to exhibit nonverbal communication within the first two years of life, such as limited eye movement and reproducible reactions to familiar people and objects. Reproducible verbal communication (sounds or specific words) was reported only for few older children. Overall, as reported by their caregivers, the queried cognitive skills are achieved by a significantly higher number of children than the queried motor skills, and only very few children are reported having unlearned a skill. No data on frequency was available for reaction to visual stimuli.

With respect to cognitive development (Figure 4, bottom), parents reported that their child exhibited consistent reactions to familiar people or objects and showed social smile in 94 %, 88 % and 85 %, respectively. Controlled eye movements were also reported for the majority of children (reaction to visual stimuli: 94 %, fixation: 88 %, eye tracking: 81 %). Reproducible communication by specific sounds or words, however, was only reported for 53 % and 17 %, respectively. Unlearned skills were only reported for a very small number of children (eye tracking and specific sounds: 2 each, fixation, social smile and specific words: 1 each). All skills were acquired considerably later than in healthy children.

### Relationship between psychomotor development and imaging findings

A clear correlation between psychomotor development, classified as “below average”, “average” and “above average” as outlined above, with brain measurements was not observed (Figure 5). The transversal cerebral diameter of most children with below-average psychomotor development was below the interpolated growth curve (Figure 5, bottom right). Similarly, above-average development in both the cognitive and motor domain was observed in children with cerebellar diameters above the interpolated growth curve (upper and lower left). No further associations between cerebral measurements and motor abilities were observed (data not shown).

**Figure 5:**
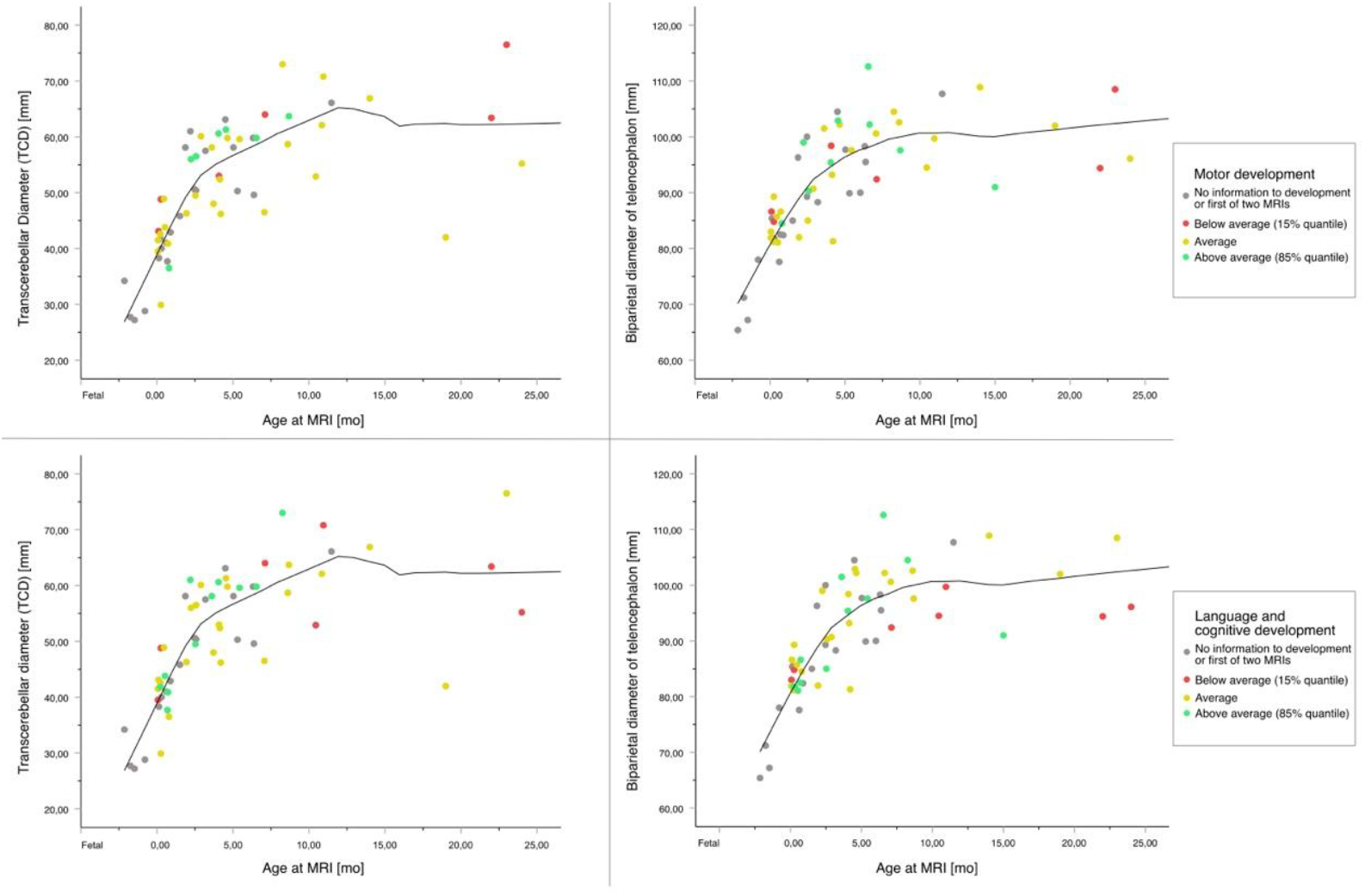
Relation of motor (upper row) and cognitive (bottom row) development with transversal diameter of the cerebellum (left) and cerebrum (right) during the first two years of life. Colors denote the classification of patients into below-average development (red), normal development (yellow) and above-average development (green). Interpolated using local estimated scatterplot smoothing (LOESS). Lower transverse cerebral diameter appears to be associated with poorer cognitive development (lower right panel), while the remaining data do not suggest meaningful correlation between brain measurements and cognitive development.

## Discussion

We analyzed brain measurements from the prenatal period into adolescence, as well as psychomotor development in a large cohort of children with PCH2A, enabling us to broaden our understanding of the imaging and clinical phenotype of this genetically very homogenous group of children.

### Growth is highly limited in both supra– and infratentorial regions

Volumetry (Figure 1) shows that differences exist between children with PCH2A and healthy controls in both cerebellar and supratentorial volume already at birth, but are more pronounced in the cerebellum. This finding is in line with a recent organoid model of PCH2a established in our group (Kagermeier et al., 2024). Healthy children then show extensive increase of supra-and infratentorial brain volumes during infancy and the following years, whereas increase of brain volumes in children with PCH2A is highly limited and stagnates after infancy. The severe growth restriction is evident in both supra– and infratentorial brain structures, but appears more pronounced and with a higher temporal dynamic in the cerebellum. In affected patients, this results in progressive microcephaly and the characteristic finding of severe cerebellar hypoplasia. Our imaging data support the clinical observation that, in early postnatal ultrasound, abnormalities are observed primarily in the cerebellum (often described as enlarged cisterna magna), whereas a majority of affected children show normal supratentorial brain structures and are still normocephalic at birth (Sánchez-Albisua et al, 2014).

### Cerebellar growth is affected in a spatial manner

Cerebellar growth is affected in a spatial manner in patients with PCH2A (Figure 2). Specifically, our measurements of the cerebellum of affected children show an increase in width and length during the first two years, but not in height, which results in the characteristic dragonfly pattern. While a slight decrease was observed in cranio-caudal height in a small number of children, the anterior-posterior and transverse diameter, as well as overall cerebellar volume, remained stable in older patients without signs of progressive degeneration in later stages of the disease. Histopathology shows both extensive dysplasia as well as ongoing atrophy of neuronal and axonal tissue, in the cerebellum of affected children (Barth et al, 2007). In cranio-caudal direction, these processes might be balanced by early growth, and could result in an apparently stable situation over time.

Alternatively, cerebellar growth might be inhibited in cranio-caudal direction right from the beginning. In the further course of the disease, processes of growth and degeneration result in a net stable cerebellar volume in older children with PCH2A. Histopathological findings suggest, that the remaining cerebellar tissue might in large parts be gliotic, rather than functional cerebellar tissue (Barth et al, 2007).

### Supratentorial structures show progressive atrophy in PCH2A

In the supratentorial structures, however, we observe ongoing tissue atrophy in children with PCH2A. The Evans index surpasses the upper threshold established for healthy children already in the first weeks of life and continues to rise over the next years (Figure 3). This index was originally established to monitor hydrocephalus and remains stable over time in healthy children (Sari et al, 2015).

However, in the absence of liquor circulation disorders or other signs of elevated intracranial pressure, it indirectly detects ventriculomegaly due to cerebral atrophy. Histological findings in the neocortex show a diffuse increase in glial tissue without active degeneration, and both cortical architecture as well as myelination of axonal tissue appeared normal even in atrophic regions (Barth et al, 2007).

In PCH2A, supratentorial atrophy takes place in a different temporal manner than cerebellar atrophy, i.e. initially less rapid, but as an ongoing process over time. Supratentorial effects of PCH2A might thus be secondary due to disrupted connections between cerebellum and neocortex, instead of a primary effect of the genetic defect (Ekert et al 2016). However, neocortical organoids derived from patients with PCH2A showed delayed growth restriction as compared to cerebellar organoids without significant signs of apoptosis in either cerebellar or neocortical organoids (Kagermaier et al, 2024).

The growth restriction of neocortical organoids was thus neither caused by cerebellar processes nor by predominant neurodegeneration, but could be a direct effect of the genetic alteration in neocortical cells. Future studies might investigate both the longitudinal neocortical neuropathology, as well as cerebro-cerebellar connections, to resolve this debate.

### Psychomotor development

Motor development (Figure 4, top) was severely delayed in all investigated patients with PCH2A. Only few patients achieved head control, attempted grasping, or were able to turn from supine to prone position. Achievement of cognitive abilities, while also delayed, was reported for a larger number of children (Figure 4, bottom). This observation corroborates clinical experience, where caregivers are frequently able to interact with their child to a certain degree, and children with PCH2A might appear limited in their expression of cognitive abilities by their motor impairments. Pronounced cerebellar pathology primarily causes motor dysfunction, given the role of the cerebellum in control and coordination of muscle activity. On the other hand, the observed discrepancy between motor and cognitive abilities of patients with PCH2A might be secondary to the questionnaire design, which assessed relatively simple cognitive skills, while the queried motor skills required a higher degree of coordination. Potential parental bias in the assessment of their child’s cognitive abilities must also be considered. Despite some histological evidence for neurodegeneration (Barth et al 2007), some developmental progress is achieved by all patients with PCH2A, and regression is relatively rare once a skill has been learned.

Regarding the individual influences of cerebellar and cerebral pathology on both motor and cognitive deficits, we found no clear relationship between morphometry and developmental trajectory. In addition to its well-known motor functions, the cerebellum has been reported to be involved in cognitive processes, and cerebellar dysfunction has been linked to cognitive impairment (Strick et al, 2009; Koziol et al, 2014; Heck et al, 2023). At the same time, the genetic defect underlying PCH2A might also affect supratentorial networks involved in both motor and cognitive development, as well as cerebro-cerebellar connections. In the present study, we focused on data that was readily obtainable from sometimes old and diverse imaging. Future studies might, however, shed more light on these questions by investigating more detailed and thorough intracranial metrics and brain networks.

### Limitations

Some limitations of this study must be considered, specifically regarding the data on developmental history. These data mostly rely on reports from caregivers, as cognitive skills might only be observed intermittently and in times of well-being or in contact with familiar people, as opposed to the unfamiliar setting of a clinical examination. While under these circumstances parental reports are an important source of information, a potentially biased perception of their child by individual caregivers cannot be ruled out. On the other hand, parental observations of their children with PCH2A fit well to the broader clinical experience.

Furthermore, clinical and imaging data were not obtained at the same time. The parental survey was completed between 2020 and 2022, while imaging data were analyzed retrospectively, and MRI were often obtained many years prior to the survey. Especially in older children with PCH2A, MRI data does not necessarily reflect the situation of the child at the time of the parental survey. This mismatch in time limits the analysis of a potential relationship between MRI and developmental parameters. It reflects the more general clinical observation that early brain MRI does not allow a precise individual prognosis regarding developmental milestones. It does not, however, affect the separate analysis of MRI metrics or psychomotor development, respectively.

## Conclusion

With the present study, we provide comprehensive data from the prenatal period to adolescence on growth patterns of intracranial structures and psychomotor development in a large cohort of children with PCH2A. Already at birth, the cerebellum of affected children was considerably smaller than in healthy children. This gap then widened progressively over the first year of life, after which cerebellar growth stagnated in children with PCH2A. Long-term data did not indicate a loss of volume in older children. In contrast, we observed ongoing supratentorial atrophy, in addition to progressive microcephaly. Caregivers reported some developmental progress, specifically in the cognitive domain, for almost all children. All motor and cognitive skills, however, were severely delayed and limited in comparison to healthy peers. Developmental regression was only reported for a limited number of children. Overall, this study expands on the previous knowledge of PCH2A both in terms of the number and the maximum age of investigated children. Insights into the long-term trajectories of this disease will not only benefit clinicians and researchers, but also the parents of affected children in knowing what to expect after such a diagnosis.

## Data Availability

All data produced in the present study are available upon reasonable request to the authors.

## Abbreviations

CI: Confidence Interval
ICC: Intraclass Correlation Coefficient
LOESS: Local Estimated Scatterplot Smoothing
mRNA: messenger RNA
PCH: Pontocerebellar hypoplasia
PCH2A: Pontocerebellar hypoplasia type 2A
tRNA: transfer RNA

## Acknowledgements

We acknowledge the kind support of the patientsߣ organization for German-speaking countries (PCH-Familie e.V.) in contacting the families of affected children.

We acknowledge support by Maximilian F Russe, Department of Diagnostic and Interventional Radiology, University Medical Center Freiburg, University of Freiburg, Freiburg, Germany.

## Author contribution statement

AH: Data acquisition, analysis, interpretation, visualization, writing – review and editing

PP: Data analysis, interpretation, visualization, writing – original draft, writing – review and editing AK: data acquisition, data interpretation

ALK: data acquisition, data interpretation

JM: Conceptualization, funding acquisition, data acquisition, writing – review and editing EK: Data analysis (NORA)

MH: statistical analysis

SM: Conceptualization, funding acquisition, supervision LL: Conceptualization, data interpretation

MU: MRI data analysis

SG: Conceptualization, funding acquisition, data interpretation, writing – review and editing

WJ: Conceptualization, funding acquisition, data acquisition, data interpretation, writing – review and editing, supervision

