## Supplementary Figures for "Brain morphometry and psychomotor development in children with PCH2A"

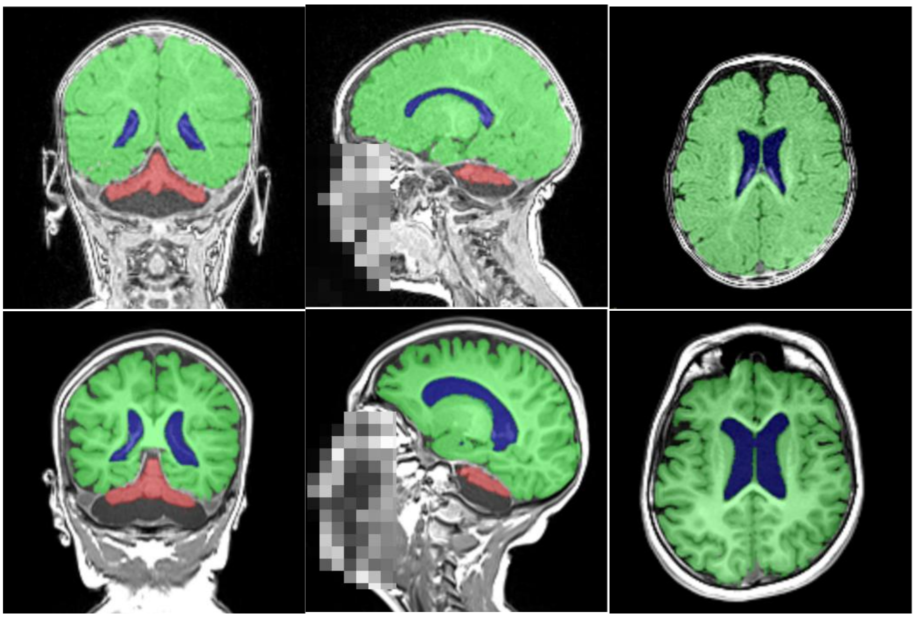

***Supplementary Figure 1:*** *Example longitudinal MRI examination and volumetric results in one child with PCH2A within the first year of life (first row) and at preschool age (second row). Note the severe cerebellar hypoplasia already in infancy as well as the increasing ventricle volume, suggesting atrophy.*

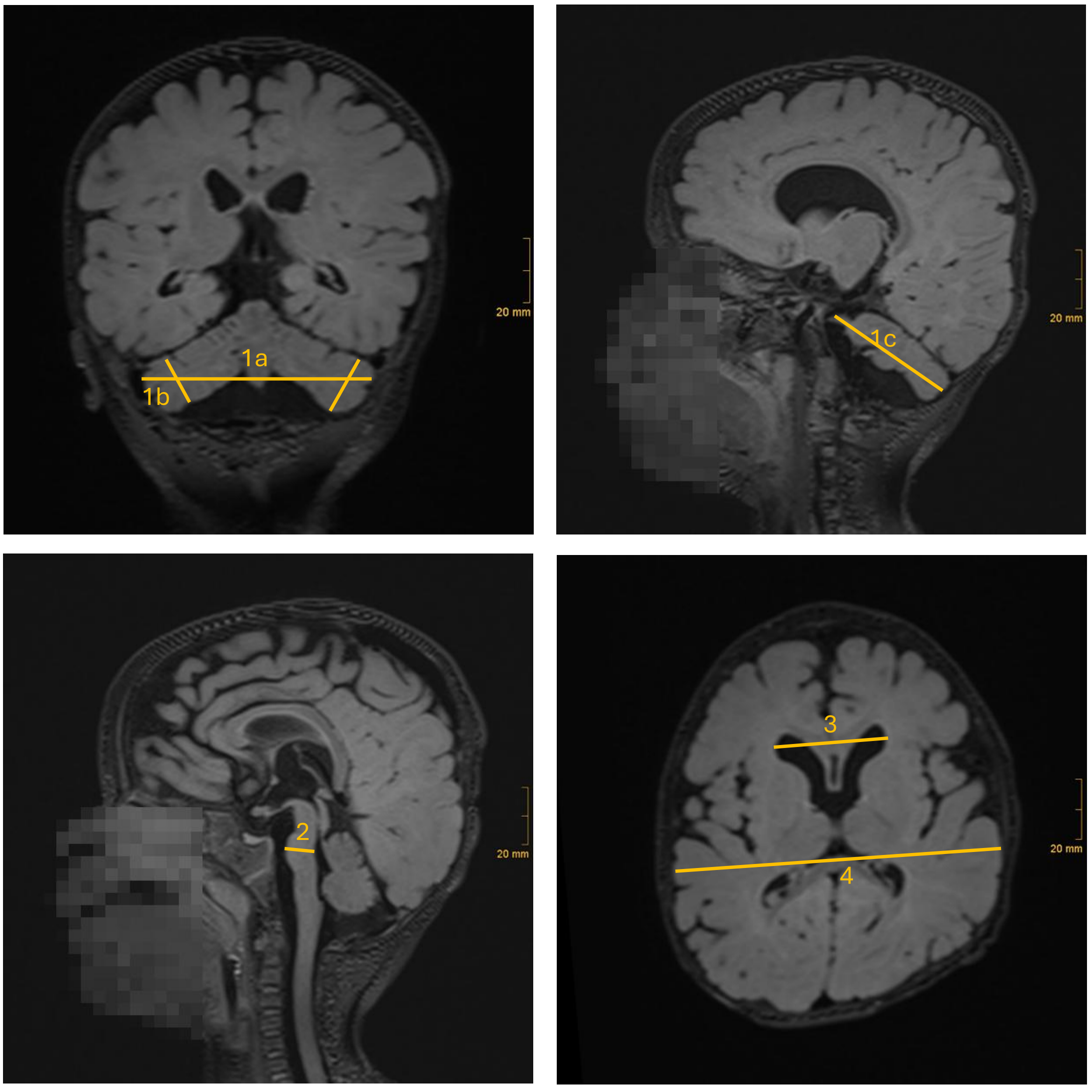


***Supplementary Figure 2:*** *Depiction of the in-plane measurements, exemplified by an MRI of a child with PCH2A in the first year of life. Upper left: Transverse (1a) and craniocaudal (1b) cerebellar diameter in the coronal plane. Upper right: anterior-posterior cerebellar length in sagittal plane (1c). Lower left: Anterior-posterior diameter of the pons in the midsagittal section (2). Lower right: Maximum diameter of the frontal horns of the lateral ventricles (3) and maximum biparietal diameter of the cerebrum in the same axial plane (4).*

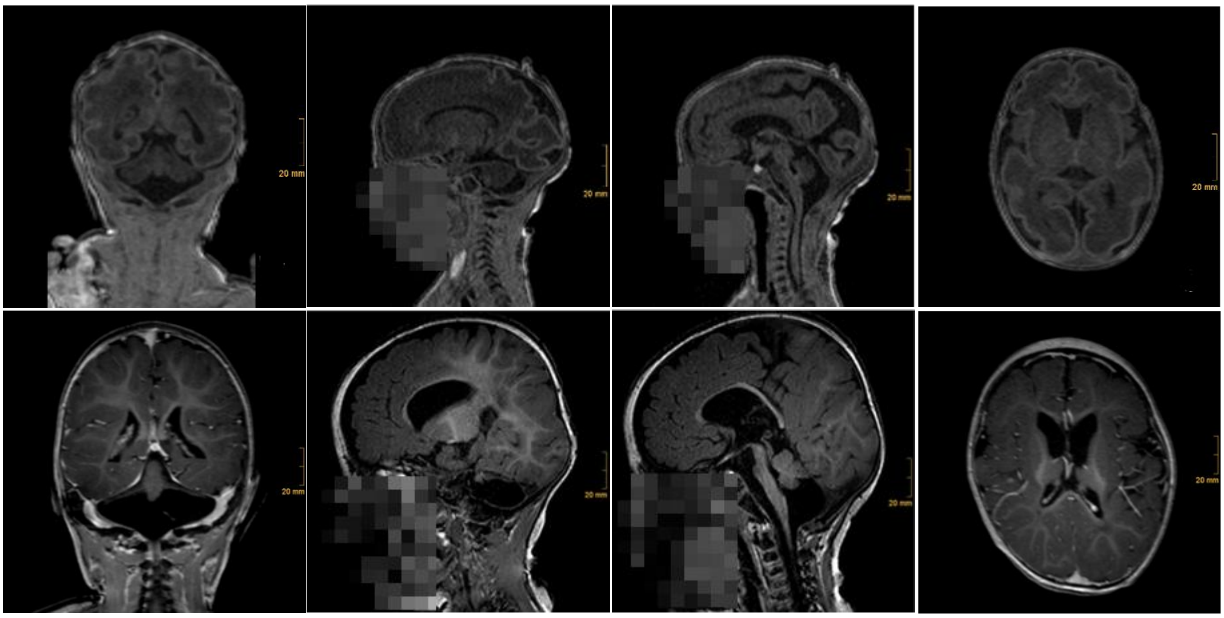


***Supplementary Figure 3:*** *Example longitudinal MRI in a very premature infant with PCH2A (<32 weeks of gestation) in the first row and less than a year later (second row). The first examination already reveals marked cerebellar hypoplasia. In the second examination, the cerebellum has grown in the transverse direction, while the craniocaudal diameter markedly decreases, potentially suggesting atrophy in this direction. Additionally, there is growth of the cerebrum in the first year alongside an increase in ventricular width.*
